# Bronchoscopy in children with diffuse alveolar hemorrhage under general anesthesia with spontaneous respiration by face mask ventilation: an analysis of thirty-eight procedures

**DOI:** 10.1101/2023.12.05.23299486

**Authors:** Ruimin Yang, Qing Wei, Xun Chen, Jing Liu, Yan Li, Jingchen Liu

**Affiliations:** Department of Anesthesiology, The First Affiliated Hospital of Guangxi Medical University, Nanning 530021, Guangxi, China; Department of Pediatrics, The First Affiliated Hospital of Guangxi Medical University, Nanning 530021, Guangxi, China

**Keywords:** bronchoscopy, diffuse alveolar hemorrhage, general anesthesia, spontaneous respiration, face mask ventilation

## Abstract

**Purpose:** To improve the management level of general anesthesia during bronchoscopy in the children with diffuse alveolar hemorrhage (DAH).

**Methods:** A retrospective study was conducted in the children with DAH who had performed bronchoscopy under general anesthesia with spontaneous respiration by face mask ventilation initially from June 2021 to June 2022 in our hospital.

**Results:** 1. Thirty-four children who had underwent 38 bronchoscopy procedures were included. 2. General anesthesia induction was performed by bolus of propofol intravenous in all the procedures. For maintaining anesthesia, 31 procedures (81.6%) received both propofol and remifentanil intravenously infusion and the rest 7 procedures (18.4%) received propofol intravenously infusion only. An intravenous bolus of ketamine or propofol was given as an anesthetic adjuvant in 21 procedures (55.3%). Thirty-five procedures (92.1%) were successfully completed under non-tracheal intubation ventilation, whereas the rest 3 procedures (7.9%) needed change to tracheal intubation ventilation. 3. Respiratory depression was found in 7 procedures (18.4%), laryngospasm was found in 2 procedure (2.6%) and bronchospasm was found in 17 procedures (44.7%). Sixteen procedures (42.1%) developed intraoperative hypoxemia. The incidence of intraoperative hypoxemia in the procedures at the active phage of disease was significantly higher compared to those at the remission phage of the disease (*P<0.05*). Sixteen procedures (42.1%) developed intraoperative hypercapnia. Two procedures (5.3%) were complicated by severe pulmonary hemorrhage.

**Conclusions:** General anesthesia with spontaneous respiration by face mask ventilation is feasible and relatively safe for the children with DAH undergoing bronchoscopy, whereas the anesthetic protocol still needs to be improved.

## Introduction

Diffuse alveolar hemorrhage (DAH) is a severe life-threatening clinical syndrome in children and can be caused by a wide variety of disorders^[1]^ . Bronchoscopy, especially bronchoalveolar lavage (BAL), transbronchial lung biopsy and cryobiopsy is crucial for the diagnosis and differential diagnosis of DAH. However, it’s a challenge for anesthesiologists to perform general anesthesia for bronchoscopy in the children with DAH, as the children with DAH usually have impaired lung function including the decreasing diffusion capacity and/or restrictive ventilation dysfunction^[2]^ . Also, some cases may have hypoxemia preoperatively^[2]^ . What’s more, the procedure of bronchoscopy may be complicated by exacerbation of pulmonary hemorrhage^[3, 4]^ . To date, a consensus is still missing over the anesthetic management of DAH in children during bronchoscopy, which can be partly explained by the relative rareness of this disease. In our hospital, total 34 children with DAH had underwent 38 bronchoscopy procedures under general anesthesia with spontaneous respiration by face mask ventilation initially from June 2021 to June 2022. Aiming to improve the management level of general anesthesia during bronchoscopy in the children with DAH, we conducted the retrospective study.

## Patients and methods

### Patients

The children aged less than 16-year-old with DAH, who were admitted in our hospital and performed bronchoscopy under general anesthesia with spontaneous respiration by face mask ventilation initially from June 2021 to June 2022 were included.

### Diagnostic criteria for DAH and definition of the disease phase

DAH was diagnosed based on respiratory symptoms (including dyspnea, hemoptysis and cough), iron deficiency anemia, diffuse pulmonary infiltrates (ground-glass opacities and/or consolidations) on the chest CT and the presence of hemosiderin laden macrophages in the BALF or lung tissue specimens^[2]^ . Active phase was defined as presence of anemia and/or diffuse pulmonary infiltrates on the chest CT and remission phase was defined as absence of hemoptysis, anemia and pulmonary infiltrates on the chest CT^[5-7]^.

### Study design

We retrospectively reviewed the medical records including the basic information, general anesthesia protocol, ventilation mode, the incidence of respiratory depression hypoxemia and hypercapnia intraoperatively and so on of the cases included in this study. The study was approved by the institutional ethics review board of the First Affiliated Hospital of Guangxi Medical University (2023-E577-01).

### Anesthesia protocol

Routinely, the children underwent a preoperative fasting period of 2 h for liquids as well as 6 h for solids and lidocaine (2%) 4 ml inhalation as well as atropine 0.01-0.02 mg/kg subcutaneously were administered 30 min before the bronchoscopy. Bronchoscopy was performed in the operating room. The children were placed in the supine position and the heart rate (HR), oxygen saturation (SpO_2_), end tidal CO_2_ (ETCO_2_) as well as blood pressure (BP) were monitored. Dexamethasone 0.1-0.3 mg/kg was injected intravenously before anesthesia induction in the cases who were in the high risk of airway spasm or postoperative nausea as well as vomiting. Bronchoscopy was performed under general anesthesia with spontaneous respiration by face mask ventilation initially, whereas tracheal intubation ventilation was well-prepared in order to ensure the safety. Firstly, general anesthesia induction was performed by bolus of propofol 2-3 mg/kg intravenous. Then, propofol 4-12 mg/kg/h was intravenously infusion for maintaining anesthesia. When needed and appropriate, remifentanil 0.1-0.2 μg/kg/min was intravenously infusion. When deepening of anesthesia was needed, an intravenous bolus of propofol 0.5 mg/kg or ketamine 0.5 mg/kg was administrated. Local anesthesia was performed by spraying of 2% lidocaine hydrochloride solution over the larynx, trachea and the main bronchi. When SpO_2_ was less than 85%, the bronchoscopy terminated. If SpO_2_ didn’t improve, pressurized ventilation by face mask or tracheal intubated ventilation if needed was performed. Also, reducing the dose of anesthetic drug was performed when respiratory depression occurred, deepening of anesthesia and dexamethasone 0.1-0.3 mg/kg or methylprednisolone 1-2 mg/kg as an intravenous bolus were performed when laryngospasm or bronchospasm occurred. Intramuscular or intravenous adrenaline was administrated in the cases with severe bronchospasm if needed.

### Definition of intraoperative respiratory depression, laryngospasm, bronchospasm, hypoxemia and hypercapnia

Intraoperative respiratory depression was defined as less than or equal to 8 breaths/min^[8]^ . Laryngospasm was defined as glottal closure accompanied by inspiratory stridor, diminished breath sounds and desaturation with SpO_2_ <92%^[9]^ . Intraoperative bronchospasm was defined as a prolonged expiratory phase accompanied by wheezing, desaturation with SpO_2_ <92% or with increasing end tidal CO_2_ (ETCO_2_) ≥50 mmHg^[9]^ . Intraoperative hypoxemia was defined as SpO_2_ <92%^[9]^ . Intraoperative hypercapnia was defined as ETCO_2_ ≥50 mmHg^[10]^.

### Statistical analysis

Statistical analysis was performed using SPSS 20.0 soft. Measurement data were expressed as medians (upper and lower range limits). Counting data were expressed as count (percentage). For the categorical variables, comparisons between groups were performed by the *χ*^2^ test or the Fisher exact test. A two-sided *P* value*<*0.05 was considered to be statistically significant.

## Results

### General information

Thirty four children (male n=7, female n=27) who had underwent 38 bronchoscopy procedures were included in this study. The median age at the time of performing bronchoscope were 85.5 months (11.0, 180.0). Thirty one procedures (81.6%) were performed at the active phage of the disease and 7 procedures (18.4%) were performed at the remission phage of the disease. Preoperative hypoxemia was found in 11 procedures (28.9%) and preoperative anemia was found in 28 procedures (73.7%).

### Anesthesia protocol and ventilation mode

General anesthesia induction was performed by bolus of propofol intravenous in all the procedures. For maintaining anesthesia, 31 procedures (81.6%) received both propofol and remifentanil intravenously infusion and the rest 7 procedures (18.4%) received propofol intravenously infusion only. An intravenous bolus of ketamine or propofol was administrated as an anesthetic adjuvant in 21 procedures (55.3%).

Of these 38 procedures, 35 procedures (92.1%) were successfully completed under the non-tracheal intubation ventilation mode, whereas the rest 3 procedures (7.9%) needed change to tracheal intubation ventilation during bronchoscopy, in which one presented with type I respiratory failure preoperatively and failed to maintain oxygenation after induction of anesthesia and the other two were complicated by severe pulmonary hemorrhage.

### Adverse events during bronchoscopy

During general anesthesia intraoperatively, respiratory depression was found in 7 procedures (18.4%), in which 6 procedures occurred when an anesthetic adjuvant was administrated by intravenous bolus of ketamine or propofol during the maintenance phase of anesthesia. Bronchospasm was found in 17 procedures (44.7%), in which 14 procedures developed bronchospasm at the time of performing BAL and 3 procedures developed bronchospasm when performing routine bronchoscopic observation. Also, 2 procedures (2.6%) developed laryngospasm, both of which occurred when the bronchoscope entered the glottis immediately following induction of anesthesia.

Sixteen procedures (42.1%) developed intraoperative hypoxemia and all of them were performed at the active phage of disease. The incidence of intraoperative hypoxemia in the procedures at the active phage of disease was significantly higher when compared to those at the remission phage of the disease (51.6% vs 0%, *P*<0.05). The most common trigger for hypoxemia was bronchospasm, which was found in 11 procedures. (Table 1)

**Table 1:** Triggers for intraoperative hypoxemia and hypercapnia.

|  | Intraoperative<br>hypoxemia(n=16) | Intraoperative<br>hypercapnia(n=16) |
| --- | --- | --- |
| Respiratory depression | 1 | 7 |
| Laryngospasm | 2 | 1 |
| Bronchospasm | 11 | 7 |
| Bleeding | 2 | 1 |

Sixteen procedures (42.1%) developed intraoperative hypercapnia, in which 10 procedures were performed at the active phage of disease and 6 procedures were performed at the remission phage of disease. The common triggers for hypercapnia were respiratory depression in 7 procedures and bronchospasm in 7 procedures. (Table 1)

In addition, two procedures (5.3%) were complicated by severe pulmonary hemorrahge, in which one occurred after BAL and another one occurred after transbronchial cryobiopsy (TBCB).

## Discussion

DAH is a clinical-pathological syndrome with accumulation of red blood cells in the alveolar spaces due to injury of the pulmonary microcirculation^[1]^ . The patients with DAH characteristically present with hemoptysis or bloody sputum, iron deficiency anemia, dyspnea and diffuse pulmonary infiltration on the chest imaging^[2]^ . Although a wide variety of disorders can result in DAH, the incidence of DAH is relatively rare in children^[11]^ . Therefore, the experience of the anesthetic management of DAH during bronchoscopy is still limited in children. In view of this, we conducted the retrospective study with the aim to improve the management level of general anesthesia during bronchoscopy in the children with DAH.

Bronchoscopy is crucial for the diagnosis and differential diagnosis of DAH. Previously, bronchoscopy under local anesthesia was widely accepted, whereas agitation was common during bronchoscopy, which not only disturbed the intraoperative manipulation, but also increased the fear of the children to the bronchoscopy. What’s more, intraoperative agitation may increase the risk of the complications, such as pneumothorax, mediastinal emphysema, aspiration and so on^[12]^ . Bronchoscopy under general anesthesia can overcome the shortcomings of the local anesthesia and have been accepted gradually by the physicians, patients and their families in recent. Whereas, the patients with DAH usually have impaired lung function and may present with hypoxemia preoperatively. Also, the procedure of bronchoscopy such as BAL, transbronchial lung biopsy and cryobiopsy may be complicated by exacerbation of pulmonary hemorrhage^[3, 4]^ . So, it’s a challenge for anesthesiologists to perform general anesthesia for bronchoscopy in the patients with DAH.

Ventilation is the safety guarantee for the bronchoscope under general anesthesia. At present, the choice of the ventilation mode during bronchoscopy is not yet uniform and is usually decided by severity of the disease, requirements of the intraoperative manipulation and the personal experience of the anesthesiologists^[13, 14]^ . In this study, all the procedures chosen non-tracheal intubation ventilation as the initial ventilation strategy and the majority were successfully completed under the non-tracheal intubation ventilation, only 3 procedures needed tracheal intubation due to the poor preoperative condition or severe pulmonary hemorrhage. This result demonstrated that non-tracheal intubation ventilation mode can be the first choice for the children with DAH for the bronchoscopy under general anesthesia, whereas intubated ventilation is required if the procedure is at the active phase of the disease with poor oxygenation preoperatively or with high risk for severe pulmonary hemorrhage.

It’s well known that reserved spontaneous breathing under general anesthesia could reduce the adverse reactions of mechanical ventilation and the length of hospital stay^[15]^ . Also, appropriate spontaneous breathing might improve gas exchange and lung aeration^[16]^ . In view of this, we attempted to perform bronchoscopy under general anesthesia with spontaneous respiration in the children with DAH in this study. Whereas, it’s a challenge to control the depth of anesthesia under general anesthesia. Excessive depth of anesthesia may result in respiratory depression and inadequate anesthesia is thought to be closely related to the airway spasm including laryngospasm as well as bronchospasm^[17, 18]^ . In this study, a propofol-based intravenous anesthesia region was administrated during bronchoscopy and the majority of the procedures were finished without severe anesthesia-related complications. However, the process was not easy, as either respiratory depress or airway spasm, especially the latter was not uncommon in this present study. Usually, pressurized ventilation by face mask or laryngeal mask was enough to cope with transient respiratory depression. In fact, some procedure bronchoscopy needed to be performed under controlled ventilation with deep anesthesia. Also, compared to respiratory depression, airway spasm seems to be trickier and not conducive to the bronchoscopy. Therefore, a deep anesthesia protocol under positive pressure ventilation seems to be preferred to bronchoscopy. As laryngeal mark is easier to be fixed and can perform control ventilation with deep anesthesia effectively, laryngeal mark seems to be superior to face mark during the bronchoscopy under general anesthesia in the children with DAH. A further study should be conduct to confirm the conclusion. Intraoperative hypoxemia is relatively common during bronchoscopy under general anesthesia^[19, 20]^ . It can be caused by laryngospasm, bronchospasm, respiratory depression, hypoventilation due to the airway obstruction, exacerbation of air-exchanging due to the introduction of saline for lavage or hemorrhage, pneumothorax and so on^[14, 19]^ . In this present study, nearly half of the procedures were complicated by intraoperative hypoxemia. Further analysis revealed that all of the procedures with intraoperative hypoxemia were performed at the active phage of disease. These results demonstrated that the poor preoperative conditions having resulted in impaired oxygenation were related to the occurrence of intraoperative hypoxemia during bronchoscopy under general anesthesia. Besides the disease itself, both bronchoscopy and anesthesia can influence the occurrence of intraoperative hypoxemia. It was worth noticing that the most common trigger for intraoperative hypoxemia during bronchoscopy under general anesthesia was bronchospasm, the occurrence of which is closely related to the proficiency of bronchoscopy practices, depth of the anesthesia and so on. Also, near half of the procedures were found to developed intraoperative hypercapnia in this study, which had been ignored by clinician in before. Unlike intraoperative hypoxemia, intraoperative hypercapnia occurred not only in the procedures at the active phage but also in those at the remission phage. Besides the bronchospasm, intraoperative hypercapnia was also related to the respiratory depression, which was mainly due to excessive anesthesia. So, it’s necessary to improve the proficiency of bronchoscopy practices and the anesthesia protocol in order to prevent or reduce the occurrence of the intraoperative hypoxemia and hypercapnia during bronchoscopy under general anesthesia.

Compared to many other illnesses, the children with DAH seem to be complicated by pulmonary hemorrhage more easily during bronchoscopy. Sequential BAL is usually performed in DAH, and the increasing red blood cell count in subsequent aliquots from the same location help to confirm the diagnosis of DAH, which is a characteristic feature for DAH^[2]^ . Negative pressure suction and/or saline mechanical irritation during the BAL may contribute to the exacerbation of the pulmonary hemorrhage^[21, 22]^ . Besides that, vascular injury due to the transbronchial biopsy or cryobiopsy is relatively common and can cause severe pulmonary hemorrhage occasionally^[4, 12, 23]^ . As reported in Koslow M et al, the incidence of severe bleeding during TBCB was 7.1% in the patients with diffuse lung disease^[23]^ . In this study, 2 procedures were complicated by severe pulmonary hemorrhage, in which one occurred after BAL and another one occurred after TBCB. And, both of the procedures needed intubation. Besides transbronchial biopsy or cryobiopsy, even the routine process like BAL may be complicated with pulmonary hemorrhage^[21, 22, 24]^ . So, it’s necessary to make up the coping strategies to prevent and/or treat the massive hemorrhage during bronchoscopy in the children with DAH and prompt tracheal intubation is needed in some cases.

This study had some limitations. Firstly, the clinical sample size was too small to apply a multivariate analysis to screen the risk factors for the complications during the bronchoscopy. Secondly, there might be misclassification or recall bias in this retrospective study. In future, a further accumulation of cases with a prospective cohort study should be conducted.

## Data Availability

All relevant data are within the manuscript and its Supporting Information files.

## Acknowledgments

This study would like to thank the participants who gave their time to contribute to this study.

## References

[1] Park JA. Treatment of Diffuse Alveolar Hemorrhage: Controlling Inflammation and Obtaining Rapid and Effective Hemostasis. Int J Mol Sci. 2021; 22:793.

[2] Lara AR, Schwarz MI. Diffuse alveolar hemorrhage. Chest. 2010; 137:1164–71.

[3] Wang S, Ye Q. Bleeding after endobronchial biopsy: sometimes frightening, often safe, always careful. BMC Pulm Med. 2023; 23:35.

[4] Herth FJF. Bronchoscopy and bleeding risk. Eur Respir Rev. 2017; 26:170052.

[5] de Prost N, Parrot A, Cuquemelle E, Picard C, Cadranel J. Immune diffuse alveolar hemorrhage: a retrospective assessment of a diagnostic scale. Lung. 2013; 191:559–63.

[6] Lichtenberger JP, 3rd, Digumarthy SR, Abbott GF, Shepard JA, Sharma A. Diffuse pulmonary hemorrhage: clues to the diagnosis. Curr Probl Diagn Radiol. 2014; 43:128–39.

[7] Spira D, Wirths S, Skowronski F, Pintoffl J, Kaufmann S, Brodoefel H, Horger M. Diffuse alveolar hemorrhage in patients with hematological malignancies: HRCT patterns of pulmonary involvement and disease course. Clin Imaging. 2013; 37:680–6.

[8] Song B, Yang Y, Teng X, Li Y, Bai W, Zhu J. Use of pre-operative anxiety score to determine the precise dose of butorphanol for intra-operative sedation under regional anesthesia: A double-blinded randomized trial. Exp Ther Med. 2019; 18:3885–92.

[9] Karaaslan E, Yildiz T. Management of anesthesia and complications in children with Tracheobronchial Foreign Body Aspiration. Pak J Med Sci. 2019; 35:1592–7.

[10] Kahl U, Yu Y, Nierhaus A, Frings D, Sensen B, Daubmann A, Kluge S, Fischer M. Cerebrovascular autoregulation and arterial carbon dioxide in patients with acute respiratory distress syndrome: a prospective observational cohort study. Ann Intensive Care. 2021; 11:47.

[11] Alexandre AT, Vale A, Gomes T. Diffuse alveolar hemorrhage: how relevant is etiology? Sarcoidosis Vasc Diffuse Lung Dis. 2019; 36:47–52.

[12] Muthu V, Ram B, Sehgal IS, Dhooria S, Prasad KT, Aggarwal AN, Agarwal R. Major complications encountered during 9979 flexible bronchoscopies performed under local anesthesia over 8 years. Lung India. 2022; 39:384–7.

[13] Abdelmalak BB, Doyle DJ. Updates and controversies in anesthesia for advanced interventional pulmonology procedures. Curr Opin Anaesthesiol. 2021; 34:455–63.

[14] Londino AV, 3rd, Jagannathan N. Anesthesia in Diagnostic and Therapeutic Pediatric Bronchoscopy. Otolaryngol Clin North Am. 2019; 52:1037–48.

[15] Lei W, Gan ZY, Liang YF, Liang CX, Jin CZ, Peng WP, Qiu XC, Guo HY. Airway foreign body caused by pepper inhalation 7 years previously retrieved under conscious sedation with spontaneous respiration: a case report. J Int Med Res. 2022; 50:3000605221086146.

[16] Chi Y, Zhao Z, Frerichs I, Long Y, He H. Prevalence and prognosis of respiratory pendelluft phenomenon in mechanically ventilated ICU patients with acute respiratory failure: a retrospective cohort study. Ann Intensive Care. 2022; 12:22.

[17] Tariq A, Hill NS, Price LL, Ismail K. Incidence and Nature of Respiratory Events in Patients Undergoing Bronchoscopy Under Conscious Sedation. J Bronchology Interv Pulmonol. 2022; 29:283–9.

[18] Jacomelli M, Margotto SS, Demarzo SE, Scordamaglio PR, Cardoso PFG, Palomino ALM, Figueiredo VR. Early complications in flexible bronchoscopy at a university hospital. J Bras Pneumol. 2020; 46:e20180125.

[19] Irmak I, Tekin F, Coplu L, Selcuk ZT. Factors related to oxygen desaturation during flexible bronchoscopy and endobronchial ultrasound. Tuberk Toraks. 2021; 69:144–52.

[20] Carlens J, Fuge J, Price T, DeLuca DS, Price M, Hansen G, Schwerk N. Complications and risk factors in pediatric bronchoscopy in a tertiary pediatric respiratory center. Pediatr Pulmonol. 2018; 53:619–27.

[21] Bada-Bosch I, Pérez-Egido L, García-Casillas MA, Del Cañizo A, Fanjul M, de la Torre M, Ordóñez J, Cerdá J, de Agustín JC. Bronchoalveolar lavage usefulness in the pediatric population. Cir Pediatr. 2020; 33:160–5.

[22] Gonski K, Cohn R, Widger J, McMullan B. Utility of bronchoscopy in immunocompromised paediatric patients: Systematic review. Paediatr Respir Rev. 2020; 34:24–34.

[23] Koslow M, Edell ES, Midthun DE, Mullon JJ, Kern RM, Nelson DR, Sakata KK, Moua T, Roden AC, Yi ES, Reisenauer JS, Decker PA, Ryu JH. Bronchoscopic Cryobiopsy and Forceps Biopsy for the Diagnostic Evaluation of Diffuse Parenchymal Lung Disease in Clinical Practice. Mayo Clin Proc Innov Qual Outcomes. 2020; 4:565–74.

[24] Anan K, Oshima Y, Ogura T, Tanabe Y, Higashi A, Iwashita Y, Fujita K, Yoshida T, Ando K, Okamori S, Okada Y. Safety and harms of bronchoalveolar lavage for acute respiratory failure: A systematic review and meta-analysis. Respir Investig. 2022; 60:68–81.

